# Off-Label Real World Experience Using Tocilizumab for Patients Hospitalized with COVID-19 Disease in a Regional Community Health System: A Case-Control Study

**DOI:** 10.1101/2020.05.14.20099234

**Authors:** Murali Ramaswamy, Praveen Mannam, Robert Comer, Emily Sinclair, D. Brent McQuaid, Monica L. Schmidt

## Abstract

**Objective:** To determine if Tocilizumab treatment in patients hospitalized with laboratory confirmed SARS-CoV-2 infection and subsequent COVID-19 disease provides short-term survival benefit.

**Design:** Case-control, observational study that includes an observation period from arrival to discharge or inpatient death. Both Cox proportional hazards and average treatment effects models were used to determine survival and treatment benefits.

**Setting:** Three Cone Health acute care hospitals including one COVID dedicated facility.

**Patients:** Patients admitted with confirmed SARS-CoV-2 from March 16, 2020 through April 22, 2020.

**Exposure:** Tocilizumab dosed at either 400 mg fixed dose or 8 mg/kg weight-based dose with maximum single dose of 800mg.

**Measurements and Main Results:** Overall, 86 patients were admitted during the observation period with confirmed COVID-19 disease. Of these, 21 received Tocilizumab during the hospital stay. Both the Cox model and treatment effects models showed short-term survival benefit. There was an associated 75% reduction in the risk of inpatient death when treated (HR 0.25; 95% CI 0.07-0.90) in the Cox model. This association was confirmed in the treatment effects model where we found a 52.7% reduced risk of dying while hospitalized compared to those not treated (RR 0.472; 95% CI 0.449-0.497). In both models, we show short-term survival benefit in patients with severe COVID-19 illness.

## Introduction

Since the emergence of SARS COV2 virus in China starting December 2019, it has evolved into a global pandemic with significant mortality and morbidity^1^. The most feared complication of COVID-19 is the development of acute respiratory distress syndrome (ARDS)^2^. It has been estimated that the mortality is in the range of 50-80% in patients with COVID-19 and respiratory failure requiring ventilation^3^.

ARDS is thought to be secondary to immune cell mediated damage of the alveolar endothelium and epithelial barrier with accumulation of protein rich edema fluid within the interstitium and alveoli^4^. There is elevation in inflammatory mediators such as Interleukin-6 (IL- 6) in the plasma of ARDS patients^5^. It appears that patients with COVID-19 have an increased inflammatory response with a cytokine storm mediated by IL-6, which is a key player in pathogenesis^6^. Elevated IL- 6 levels are associated with worse outcomes in patients with COVID-19^7^. Given the central role of IL-6 in the pathogenesis of COVID-19 infection and ARDS there is an increased interest in testing IL-6 inhibition during treatment in severely ill COVID-19 patients.

Multiple reports in the course of this pandemic strongly suggest a potential role for IL-6 inhibition in reducing mortality or improving clinical outcomes^7-10^. However, many of these studies lack a control arm^9^. Despite the Food and Drug Administration’s (FDA) Emergency Use Approval (EUA) for the antiviral Remdesivir, there is a strong unmet need for reduction of ARDS related COVID-19 mortality and morbidity via the inhibition of cytokine storm.

Tocilizumab is one of several immunomodulating monoclonal antibodies targeting the IL-6 pathway. It was developed to treat rheumatoid arthritis (RA) and systemic juvenile idiopathic arthritis^11^. Tocilizumab competitively inhibits the binding of IL-6 to both its soluble and membrane-bound receptors. By inhibiting the receptor complex, IL-6 signal transduction to inflammatory mediators that summon B and T cells is halted^12^.

Cone Health is a regional health system with multiple centers of excellence and over 800 inpatient beds across six hospitals that includes a dedicated COVID-19 inpatient facility. In response to the COVID-19 pandemic, the Cone Health Antimicrobial Program ("CHAMP”) issued a treatment algorithm guideline for the bedside clinician. The first version was issued on March 24, 2020 based on the first published literature from China^9^. This included Tocilizumab use on an off-label basis restricted to critically ill ventilated patients. In this version, Tocilizumab dosing was set at 400mg IV single fixed dose. Due to an influx of emerging data, the treatment algorithm was updated on April 3, 2020 to allow all patients with a positive test and an oxygen saturation <= 88% concomitant with evidence of cytokine storm (C-Reactive Protein >= 7 mg/dL) to be considered for a dose of 400 mg of tocilizumab^13^. On April 13, 2020 the algorithm was further updated with the introduction of weight based dosing and exclusion criteria to better reflect the ongoing COVACTA trial^13^. These changes and updates were communicated to all prescribing providers via email. Given the off-label indication, the decision to prescribe Tocilizumab was left to the individual treating physician.

This study aims to evaluate the impact Tocilizumab has on short-term survival in patients with severe COVID-19 infection. Tocilizumab is currently being evaluated in a large randomized, double-blind, placebo controlled phase III trial (COVACTA, NCT04320615, Sponsor: Hoffmann-La Roche) to assess the efficacy of the drug in patients with severe COVID-19 pneumonia^13^. Our study presents an early, observational evaluation of this drug and the association with inpatient mortality.

## Methods

### Study design and Setting

This is an observational case-control study design. Cases were patients that died while hospitalized while controls were discharged alive. The exposure was treatment with Tocilizumab during the inpatient stay. Data were analyzed using Stata statistical software version 16.1 (Statacorp, College Station, TX). Data were obtained through Cone Health’s Enterprise Analytics data warehouse where data are sourced from our electronic medical record (Epic Systems). This study received approval from Cone Health’s institutional review board as exempt from full review (IRB#1604456-1).

## Outcome

The primary endpoint for the study was inpatient mortality. We analyzed eighty-six consecutive patients that were hospitalized with laboratory confirmed SARS-CoV-2 infection using nucleic acid amplification testing (**Figure 1**). The observation period commenced on March 16, 2020, the date of the first hospitalization for COVID-19 in the health system, and ended with the last COVID-19 hospitalization on April 22, 2020. The cut-off date was arbitrarily chosen for preparation of the manuscript. Twenty-one patients were treated with Tocilizumab. A 400mg fixed dose was administered to fourteen patients prior to switching to an 8 mg/kg dosing strategy April 13, 2020. A total of seven patients received weight-based dosing with a maximum dose of 800 mg.

**Figure 1:**
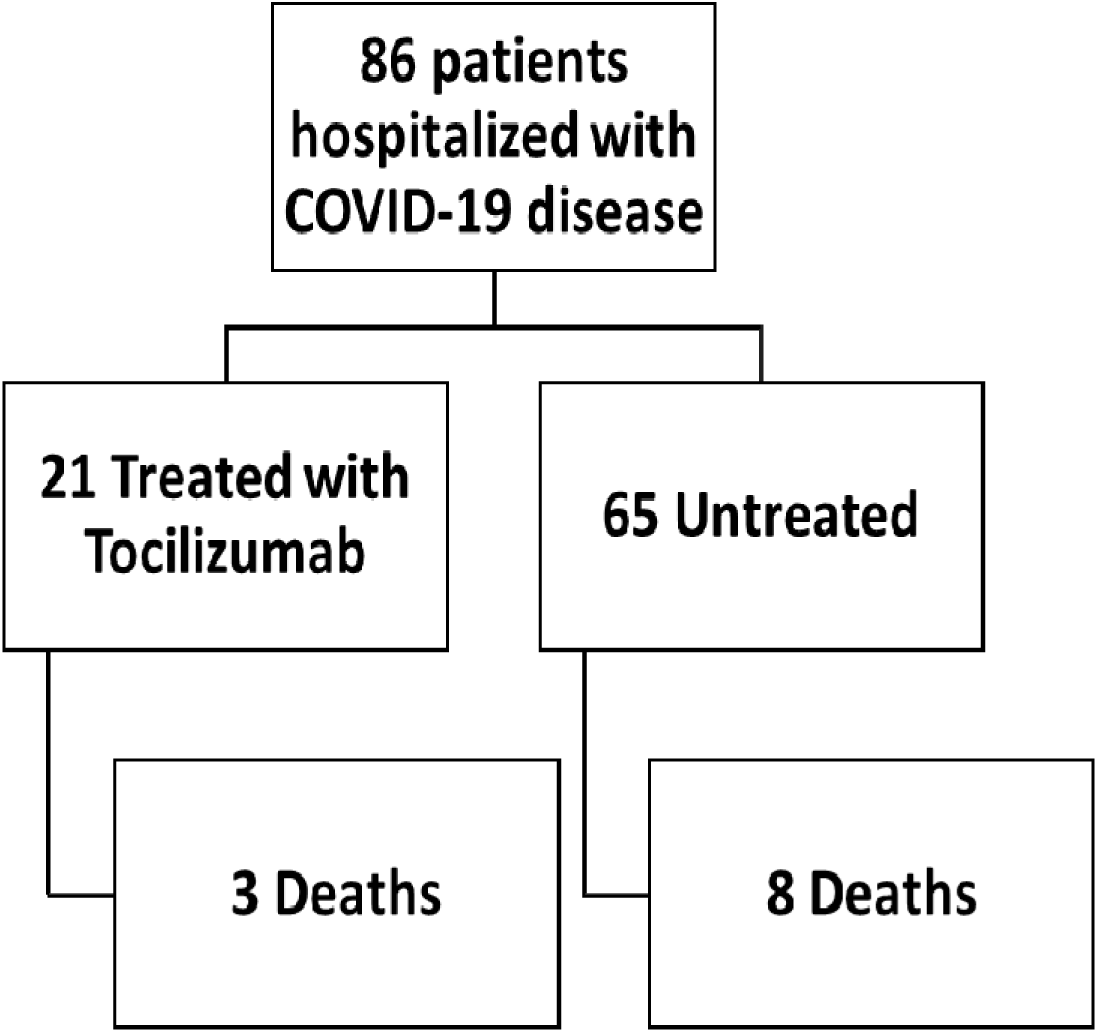
CONSORT Diagram

## Statistical Analyses

Three analyses were used to test our hypothesis that Tocilizumab is associated with a decrease in the risk in inpatient mortality and shows a short-term survival benefit. First, we compared demographics, clinical presentation, comorbidity burden and socioeconomic factors between those treated versus those that were untreated without adjustment (univariate). We used t-tests for continuous variables and Chi-square for binary and categorical variables^14^. Second, we analyzed the association of treatment with short-term survival using a Cox proportional hazards model^15^. The Cox model was limited to the number of covariates that could be included in order to avoid violation of the proportional hazards assumption. We therefore analyzed a more robust logit model to determine average treatment effects while using inverse probability weighting to adjust for treatment selection bias^16^. Both the survival and average treatment effects models are shown and discussed.

### Univariate

We compared enrolled patients’ characteristics by treatment status (**Table 1**). We used t-tests for continuous variables and Chi-square for binary or categorical variables.

**Table 1:** Univariate comparisons between those treated with Tocilizumab and untreated.

| **Clinical Variable** | **Overall**<br>**n=86** | **Untreated**<br>**n=65** | **Treated**<br>**n=21** | **p-value** |
| --- | --- | --- | --- | --- |
| **Inpatient Death (%)** | 12.8 [11] | 12.3 [8] | 14.3 [3] | 0.81 |
| **Nasal cannula flow rate >=6L (%)** | 26.7 [23] | 20 [13] | 47.6 [10] | 0.01* |
| **Average Age at Admission –years (SD)** | 63.7<br>(15.7) | 63.8 (15.9) | 63.2 (15.6) | 0.87 |
| **<u>Tocilizumab (# patients)</u>** |  |  |  |  |
| **Fixed dosing regimen 400 mg** | N/A | N/A | 14 |  |
| **Weight based dosing (8mg/kg)** | N/A | N/A | 7 |  |
| **Patients receiving 2 doses of**<br>**Tocilizumab** | N/A | N/A | 8 |  |
| **Tocilizumab administered prior to**<br>**mechanical ventilation** | N/A | N/A | 10 |  |
| **Tocilizumab administered after**<br>**mechanical ventilation** | N/A | N/A | 11 |  |
| **Treated patients ever requiring**<br>**mechanical ventilation** | N/A | N/A | 12 |  |
| **<u>Race (%)</u>** |  |  |  | 0.33 |
| **White** | 44.2 [38] | 41.5 [27] | 52.4 [11] |  |
| **Black** | 46.5 [40] | 50.8 [33] | 33.3 [7] |  |
| **Other** | 9.3 [8] | 7.7 [5] | 14.3 [3] |  |
| **<u>Gender</u>** |  |  |  |  |
| **Male** | 57 [49] | 55.4 [36] | 61.9 [13] | 0.6 |
| **Female** | 43 [37] | 44.6 [29] | 38.1 [8] |  |
| **<u>Marital Status</u>** |  |  |  |  |
| **Married/Partner/Significant other** | 61.6 [53] | 61.5 [40] | 61.9 [13] | 0.85 |
| **Divorced/Separated/Widowed** | 12.8 [11] | 13.8 [9] | 9.5 [2] |  |
| **Other/single** | 25.6 [22] | 24.6 [16] | 28.6 [6] |  |
| **Average Elixhauser Comorbidity Score**<br>**(SD)** | 2.6 (4.3) | 2.6 (4.4) | 2.6 (4.0) | 0.98 |
| **Average Body Mass Index** | 33.5<br>(8.3) | 33.8<br>(8.4) | 32.8<br>(8.2) | 0.66 |
| **History of Diabetes (%)** | 11.6 [10] | 10.8 [7] | 14.3 [3] | 0.66 |
| **History of COPD (%)** | 26.7 [23] | 26.2 [17] | 28.6 [6] | 0.83 |
| **Current or former smoker (%)** | 3.5 [3] | 3.1 [2] | 4.8 [1] | 0.71 |
| **History of Hypertension (%)** | 20.9 [18] | 23.1 [15] | 14.3 [3] | 0.39 |
| **History of Heart Disease (%)** | 4.7 [4] | 4.6 [3] | 4.8 [1] | 0.98 |
| **History of Cancer (%)** | 2.3 [2] | 3.1 [2] | 0.0 [0] | 0.42 |
| **History of Atrial Fibrillation (%)** | 7.0 [6] | 6.2 [4] | 9.5 [2] | 0.60 |
| **History of Stroke (%)** | 2.3 [2] | 1.5 [1] | 4.8 [1] | 0.39 |
| **History of Anemia (%)** | 5.8 [5] | 7.7 [5] | 0.0 [0] | 0.19 |
| **History of Peripheral Vascular Disease (%)** | 2.3 [2] | 1.2 [1] | 4.8 [1] | 0.39 |
| **Average MEWS Score during the stay (SD)** | 1.4 (1.6) | 1.1 (1.3) | 2.5 (1.9) | <0.001* |
| **Placed on HFNC (%)** | 22.1 [19] | 16.9 [11] | 38.1 [8] | 0.04* |
| **Maximum Nasal Cannula Flow Rate during the stay -Liters(SD)** | 5.0 (7.6) | 4.7 (8.2) | 5.9 (5.4) | 0.55 |
| **<u>Level of Care (%)</u>** |  |  |  |  |
| **ICU** | 20.9 [18] | 12.3 [8] | 47.6 [10] | <0.001* |
| **Required Mechanical Ventilation (%)** | 26.7 [23] | 15.4 [10] | 61.9 [13] | <0.001* |
| **<u>Prone Status (%)</u>** |  |  |  |  |
| **Prone and Mechanical Ventilation** | 8.1 [7] | 1.5 [1] | 28.6 [6] | <0.001* |
| **Prone and No Mechanical Ventilation** | 20.9 [18] | 20.0 [13] | 23.8 [5] |  |
| **Not prone and Mechanical Ventilation** | 18.6 [16] | 13.8 [9] | 33.3 [7] |  |
| **Not prone and No Mechanical Ventilation** | 52.3 [45] | 64.6 [42] | 14.3 [3] |  |
| **Opioid medication doses during the stay (SD)** | 4.6 (7.3) | 3.2 (6.3) | 9.0 (8.8) | 0.001* |
| **Anti-Diabetes medication doses during the stay (SD)** | 3.5 (6.4) | 2.9 (5.1) | 5.4 (9.1) | 0.13 |
| **Vasopressor doses during the stay (SD)** | 0.9 (1.9) | 0.7 (1.9) | 1.6 (1.8) | 0.09 |
| **Anticoagulant medication doses during the stay (SD)** | 2.9 (4.2) | 2.4 (4.4) | 4.4 (3.2) | 0.06 |
| **Antibiotic doses during the stay (SD)** | 4.1 (3.2) | 4.0 (3.1) | 4.4 (3.4) | 0.59 |
| **<u>Concomitant medications used during the stay(%)</u>** |  |  |  |  |
| **Azithromycin** | 32.6 [28] | 35.4 [23] | 23.8 [5] | 0.33 |
| **Hydroxychloroquine** | 67.4 [58] | 63.1 [41] | 81.0 [17] | 0.13 |
| **ACE Inhibitors** | 10.5 [9] | 9.2 [6] | 14.3 [3] | 0.51 |
| **Corticosteroids** | 20.9 [18] | 13.8 [9] | 42.9 [9] | 0.004* |
| **Received Continuous Renal Replacement Therapy (%)** | 9.3 [8] | 4.6 [3] | 23.8 [5] | 0.008* |

| **<u>Laboratory Results \*\*</u>** |  |  |  |  |
| --- | --- | --- | --- | --- |
| **AST U/L** | 55.2<br>(37.9)<br>[82] | 53.5 (32.7)<br>[61] | 60.4 (50.6)<br>[21] | 0.47 |
| **ALT U/L** | 38.9<br>(32.5)<br>[82] | 37.3 (29.1)<br>[61] | 43.5 (41.6)<br>[21] | 0.45 |
| **C-Reactive Protein mg/dL** | 12.6 (8.3)<br>[62] | 11.2 (8.5)<br>[44] | 15.9 (6.9)<br>[18] | 0.05* |
| **IL-6 pg/mL** | 211.5<br>(339.0)<br>[23] | 64.4 (45.9)<br>[12] | 371.9<br>(443.0) [11] | 0.03* |
| **D-Dimer ug/mL-FEU** | 2.3 (2.3)<br>[45] | 2.0 (2.0)<br>[30] | 2.9 (2.7)<br>[15] | 0.19 |
| **Glucose mg/dL** | 161.4<br>(83.3)<br>[86] | 157.2<br>(85.7)<br>[65] | 174.6<br>(76.1)<br>[21] | 0.41 |
| **Estimated GFR** | 39.8<br>(19.0)<br>[43] | 38.8 (19.0)<br>[33] | 42.9 (20.0)<br>[10] | 0.56 |
| **<u>Quantiferon TB-Gold Plus (%)</u>** |  |  |  |  |
| **Indeterminate** | 5.8 [5] | 1.5 [1] | 19.0 [4] | <0.001* |
| **Negative** | 7.0 [6] | 0.0 [0] | 28.6 [6] |  |
| **Positive** | 2.3 [2] | 1.5 [1] | 4.8 [1] |  |
| **Not tested** | 84.9 [73] | 96.9 [63] | 47.6 [10] |  |
| **Absolute Lymphocyte count (SD)** | 1.0 (0.5)<br>[84] | 1.0 (0.5)<br>[63] | 1.1 (0.6)<br>[21] | 0.53 |
| **Absolute Neutrophil count (SD)** | 5.6 (3.1)<br>[84] | 5.3 (3.3)<br>[63] | 6.7 (2.5)<br>[21] | 0.08 |
| **Platelet count K/uL** | 203.6<br>(77.9)<br>[84] | 204.6<br>(81.5)<br>[63] | 200.5<br>(67.9)<br>[21] | 0.84 |
| **Procalcitonin ng/mL** | 1.6 (3.8)<br>[40] | 1.4 (3.7)<br>[30] | 2.2 (4.0)<br>[10] | 0.56 |
| **INR** | 1.4 (1.0)<br>[26] | 1.3 (0.4)<br>[13] | 1.6 (1.4)<br>[13] | 0.47 |
| **Ferritin ng/mL** | 846.1<br>(1158.6)<br>[58] | 784.5<br>(1275.3)<br>[40] | 982.9<br>(860.4)<br>[18] | 0.55 |
| **Serum Creatinine mg/dL** | 1.8 (2.3)<br>[86] | 1.9 (2.3)<br>[65] | 1.7 (2.2)<br>[21] | 0.8 |
| **Blood urea nitrogen mg/dL** | 24.9<br>(22.7)<br>[84] | 25.2 (24.4)<br>[64] | 24.0 (16.7)<br>[20] | 0.83 |
| **Total bilirubin mg/dL** | 0.9 (0.4)<br>[82] | 0.9 (0.4)<br>[61] | 0.8 (0.4)<br>[21] | 0.66 |
| **Alkaline phosphatase U/L** | 74.1<br>(35.2)<br>[82] | 72.9 (37.0)<br>[61] | 77.6 (29.6)<br>[21] | 0.6 |
| **Average SOFA score during the stay** | 2.6 (4.0)<br>[29] | 2.1 (3.6)<br>[19] | 4.0 (4.9)<br>[10] | 0.06 |
| **Patients=[n]** |  |  |  |  |
| **\*Statistical significance threshold: $p \leq 0.05$** |  |  |  |  |
| **\*\*First laboratory value resulted after admission-units (Standard Deviation)[n=patients with results]** |  |  |  |  |
| **P-values by t-test for continuous variables and chi2 test for binary/categorical variables** |  |  |  |  |

### Multivariate

We used Cox proportional hazards with robust Huber-White standard errors to evaluate the association between Tocilizumab and survival to discharge^17^. Patients were considered under observation from arrival to the emergency department or inpatient room for outside transfer to death or discharge. Censoring occurred at discharge. Observation time was measured in days and there were 474.5 total days included in the at-risk analysis time. Failure was inpatient death. There were 11 failures during the observation period (3 in the treated and 8 in the untreated). The Cox proportional hazards assumption was not violated (p=0.3477).

The model adjusted for the following: age at admission, race, gender, marital status, Elixhauser-van Walraven comorbidity score, average modified early warning score (MEWS) during the inpatient stay, use of high-flow nasal cannula, maximum flow rate used on nasal cannula, highest level of care (ICU, Progressive, etc.), if mechanical ventilation was required, number of opioid doses, number of anti-diabetic drug doses, number of anticoagulant doses and number of antibiotic doses. Drugs were grouped by drug class. Laboratory values were frequently missing and are not included in the multivariate adjustment. We report hazard ratios from the Cox model (**Table 2**), smoothed hazard curves (**Figure 2**) and Cox model adjusted survival curves (**Figure3**). We were not able to include all covariates of interest due to violation of the proportional hazards assumption. We support our findings by including a more robust average treatment effects logit model with inverse probability weighting (**Table 3**).

**Table 2:** Cox Proportional Hazard Model

| **Covariate** | **Hazard Ratio** | **Robust SD** | **Z** | **95% CI** |  | **p-value** |
| --- | --- | --- | --- | --- | --- | --- |
|  |  |  |  | **Low** | **High** |  |
| **Treated(Tocilizumab)** | 0.25 | 0.16 | -2.12 | 0.07 | 0.90 | 0.034* |
| **Age at Admission** | 1.07 | 0.02 | 2.91 | 1.02 | 1.12 | 0.004* |
| **Race** | 1.03 | 0.51 | 0.06 | 0.39 | 2.74 | 0.95 |
| **Gender** | 0.51 | 0.57 | -0.61 | 0.06 | 4.55 | 0.544 |
| **First serum creatinine post admission** | 1.01 | 0.27 | 0.03 | 0.59 | 1.72 | 0.973 |
| **Elixhauser comorbidity score** | 1.05 | 0.16 | 0.33 | 0.78 | 1.43 | 0.738 |
| **Average MEWS** | 2.44 | 0.86 | 2.54 | 1.23 | 4.85 | 0.011 |
| **Received HFNC** | 0.47 | 0.55 | -0.64 | 0.05 | 4.79 | 0.521 |
| **Highest flow rate (nasal cannula)** | 1.06 | 0.05 | 1.46 | 0.98 | 1.16 | 0.143 |
| **Level of Care** | 1.05 | 0.61 | 0.09 | 0.34 | 3.25 | 0.927 |
| **Required mechanical ventilation** | 15.73 | 40.88 | 1.06 | 0.10 | 2565.35 | 0.289 |
| **Opioid doses** | 0.78 | 0.07 | -2.92 | 0.66 | 0.92 | 0.004* |
| **Anti-diabetes doses** | 0.87 | 0.11 | -1.13 | 0.69 | 1.11 | 0.258 |
| **Anti-coagulant doses** | 0.86 | 0.08 | -1.64 | 0.71 | 1.03 | 0.101 |
| **Antibiotic doses** | 1.00 | 0.10 | 0.01 | 0.82 | 1.23 | 0.992 |

**Figure 2:**
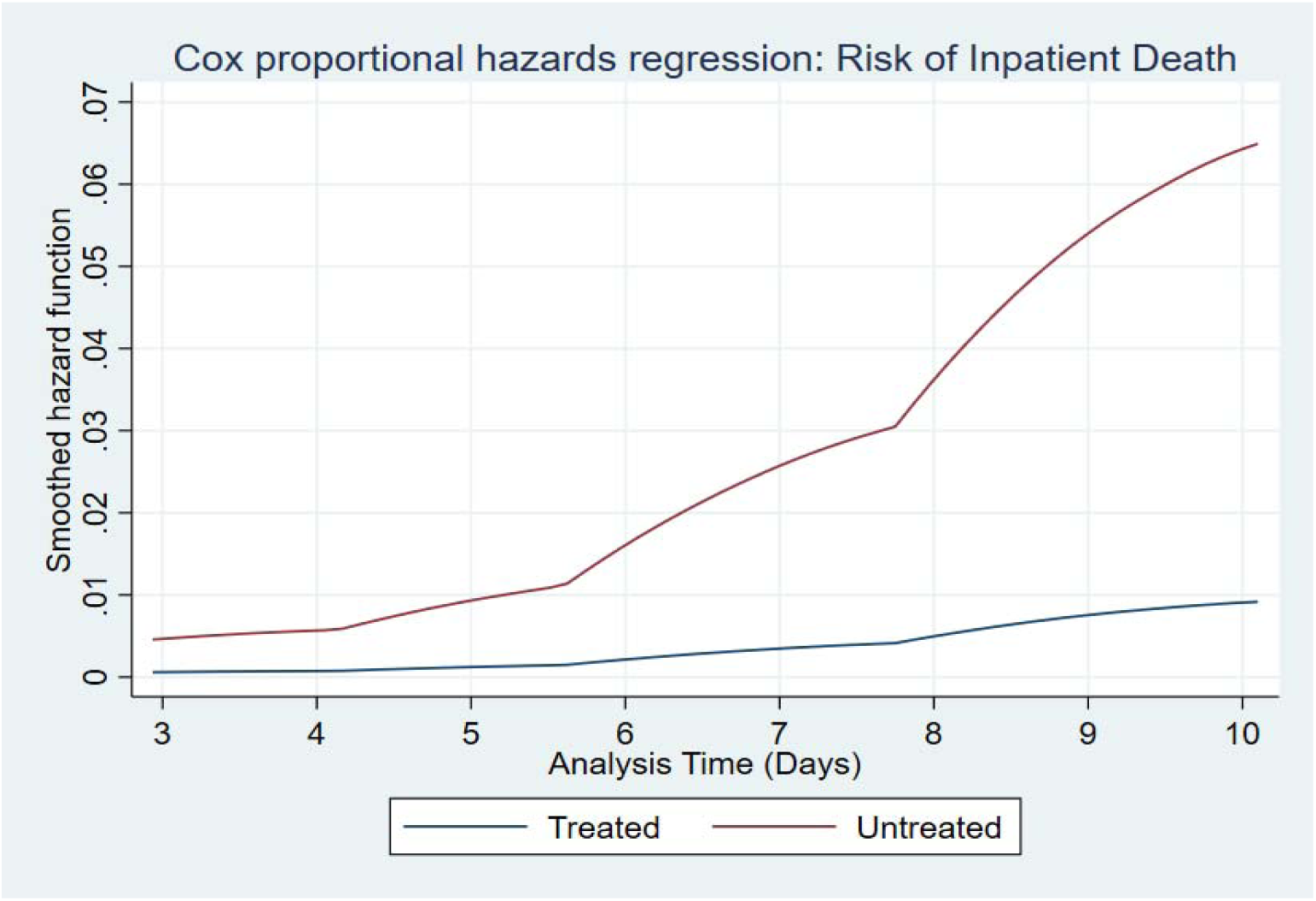
Smoothed Hazard Function. *During the observation time, patients treated with Tocilizumab have statistically significant lower hazard of inpatient death compared to those not treated*.

**Figure 3:**
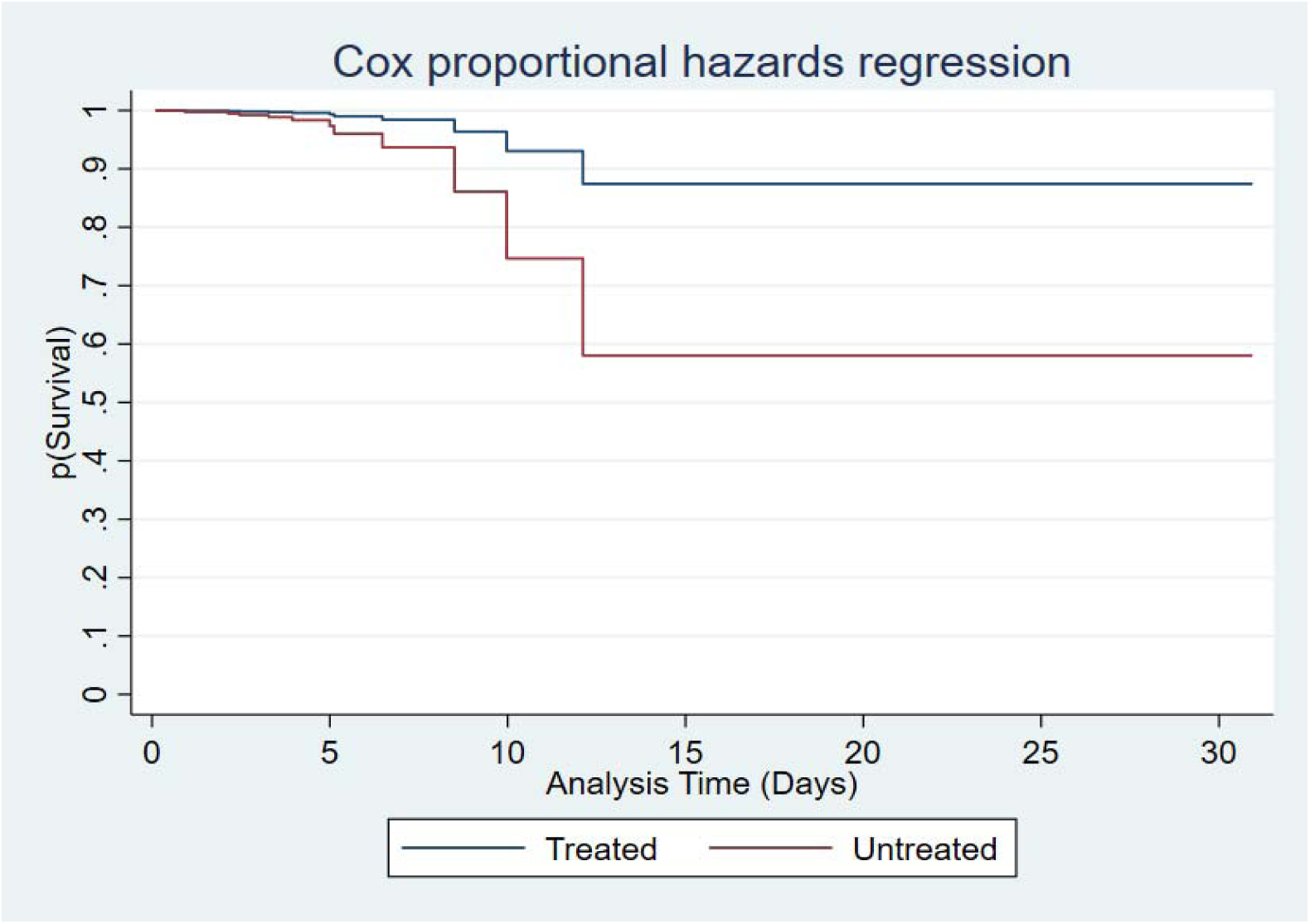
Adjusted Cox Proportional Hazards Survival Curves. *For patients not treated with Tocilizumab, the probability of survival over the observation period is significantly less than the treated when adjusting for competing risk of mortality*.

**Table 3:** Average Treatment Effects Model with Inverse Probability Weights

|  | Relative Risk | Robust SE | Z | 95% CI |  | p-value |
| --- | --- | --- | --- | --- | --- | --- |
|  |  |  |  | Low | High |  |
| **Average Treatment Effect for Population (Treated vs. Untreated)** | 0.473 | 0.498 | -2.2 | 0.449 | 0.497 | 0.028* |
| **Average Population Outcome Mean** | 0.533 | 0.038 | 3.49 | 0.514 | 0.552 | <0.001* |
| **<u>Covariates</u>** |  |  |  |  |  |  |
| **Age at admission** | 0.50 | 0.02 | -0.41 | 0.486 | 0.509 | 0.68 |
| **Race** | 0.36 | 0.49 | -1.22 | 0.176 | 0.589 | 0.22 |
| **Gender** | 0.39 | 0.80 | -0.56 | 0.118 | 0.754 | 0.58 |
| **Marital status** | 0.47 | 0.41 | -0.33 | 0.279 | 0.663 | 0.74 |
| **Elixhauser Score** | 0.52 | 0.09 | 0.91 | 0.476 | 0.565 | 0.36 |
| **Average MEWS during stay** | 0.50 | 0.29 | 0.06 | 0.365 | 0.644 | 0.95 |
| **Highest flow rate (nasal cannula)** | 0.49 | 0.05 | -0.52 | 0.466 | 0.520 | 0.60 |
| **Received high-flow nasal cannula treatment** | 0.61 | 0.92 | 0.49 | 0.206 | 0.905 | 0.63 |
| **Received mechanical ventilation** | 0.91 | 1.16 | 1.97 | 0.504 | 0.990 | 0.048* |
| **Patient placed in prone position during the stay** | 0.89 | 0.85 | 2.48 | 0.609 | 0.978 | 0.01* |
| **Received continuous renal replacement therapy** | 0.93 | 2.05 | 1.25 | 0.189 | 0.999 | 0.21 |
| **Antibiotics Doses during stay** | 0.45 | 0.14 | -1.49 | 0.382 | 0.516 | 0.14 |
| **First Serum Creatinine at admission** | 0.41 | 0.35 | -1.07 | 0.257 | 0.577 | 0.28 |
| **Other COVID medications (Azithromycin/Hydroxychlorquine/Corticosteroids)** | 0.79 | 2.10 | 0.64 | 0.060 | 0.996 | 0.52 |
| **Model Constant** | 0.22 | 1.73 | -0.75 | 0.009 | 0.890 | 0.46 |

Covariates for the Cox and treatment effects models were chosen based on factors believed to be associated with inpatient mortality. In order to elucidate the effect of Tocilizumab, we adjusted for patient-level demographics and socioeconomic factors. To adjust for the patient’s comorbidity burden, we used the Elixhauser-van Walraven comorbidity score that has been validated as a predictor of inpatient mortality^18-20^. The Elixhauser-von Walraven score includes historical documentation of the following; congestive heart failure, cardiac arrhythmias, valvular disease, pulmonary circulation disorders, peripheral vascular disorders, hypertension, neurodegenerative disorders, chronic pulmonary disease, diabetes, hypothyroidism, renal failure, liver disease, non-bleeding peptic ulcer, AIDS/HIV, lymphoma, metastatic cancer, solid tumor without metastasis, rheumatoid arthritis/collagen diseases, coagulopathy, obesity, weight loss, fluid and electrolyte disorders, blood loss anemia, deficiency anemia, alcohol abuse, drug abuse, psychosis and depression. Anti-diabetic, opioid and anti-coagulant drug dose counts during the encounter were added to the Cox model as a proxy of severity of illness and stability. For example, patients with a glucose that fluctuates frequently may require more insulin to maintain control and indicate the patient is not as stable as someone with fewer or no insulin doses.

### Average Treatment Effects Model

This model allows estimation of the effect of Tocilizumab on the risk of inpatient mortality independent of the decision to treat. Using inverse probability weights that adjust estimates based on the probability a patient received the drug helps mitigate selection bias and offers robust estimates. We used a logit model with inverse probability weighting and robust Huber-White standard errors. The estimates were transformed to relative risk using the following formula:

#### exp(estimate)/1+exp(estimate)

This model allowed inclusion of more covariates without violating the overlap assumption. We adjusted for all patient demographics, socioeconomic factors, clinical factors and were able to include average serum creatinine, azithromycin, hydroxychloroquine, corticosteroids, continuous renal replacement therapy and prone positioning (Table 3).

## Results

In the univariate analyses, our study found no statistically significant difference in baseline comorbidity burden (Elixhauser-von Walraven comorbidity index) between those treated with and those not treated with Tocilizumab. The average SOFA scores over the length of the admission also did not differ between the two groups. However, treated patients displayed higher levels of biomarkers indicative of cytokine storm (CRP and IL-6) at initial presentation. In addition, they displayed greater physiologic instability during the course of their stay as evidenced by greater respiratory distress burden as evidenced by higher levels of oxygen need and mechanical ventilation, and a higher need for acute renal replacement therapy. Specifically, we found that where laboratory data was available, the pre-treatment C-reactive protein (11.2 vs. 15.9 mg/dL) and IL-6 (64.4 vs. 371.9 pg/mL) values were higher in the treated compared to untreated patients. Additionally, in those treated with Tocilizumab, the average MEWS score was 1.4 points higher than the untreated (p<0.001). Furthermore, 47.6% of those in the Tocilizumab treatment group required six liters or more of oxygen by nasal cannula compared to only 20% of the untreated. The majority of patients in the treatment group received mechanical ventilation compared to those not treated (61.9% vs. 15.4%). Continuous renal replacement therapy (23.8% vs. 4.6%) and corticosteroid use (42.9% vs. 13.8%) was more frequent in the treated compared to untreated respectively.

The adjusted Cox proportional hazards model showed treatment with Tocilizumab was associated with an 75% reduced risk of dying while hospitalized compared to those not treated (HR 0.25; 95% CI 0.07-0.90) (**Table 2**). This association was robust to adjustment for clinical and patient-level factors that may impact the risk of dying while hospitalized (Figure 2). For each additional point gained in the modified early warning score (MEWS), the risk of death increased by 2-fold (HR 2.44; 95% CI 1.23-4.85). The Cox proportional hazards survival curves show probability of survival at specific days during a patient’s inpatient stay by treatment group (**Figure 3**). Patients censor out if discharged.

The treatment effects model indicated that patients treated with Tocilizumab have an associated 52.7% reduced risk of dying while hospitalized compared to those not treated (RR 0.472; 95% CI 0.449-0.497) (**Table 3**). Because the treatment effects model evaluates the counterfactual, we are also able to conclude that the risk of dying while hospitalized falls by 81.4% (Estimate −0.814; 95% CI −1.38, −0.247) if everyone in our sample were treated with Tocilizumab compared to if none were treated.

Taken together, our data indicate that bedside physicians chose to prescribe Tocilizumab to a population of patients displaying more severe illness and acute lung Injury as evidenced by MEWS score, cytokine biomarkers, oxygen dependence, mechanical ventilation and renal replacement therapy. Nevertheless, the decision to prescribe Tocilizumab in this population was associated with lower risk of short-term mortality.

## Discussion

Our analysis suggest that Tocilizumab may offer short-term survival benefit in patients with more severe COVID-19 disease. Tocilizumab is currently approved to treat conditions with cytokine release syndrome such as chimeric antigen receptor (CAR) T-cell therapy and is proven to reduce cytokine mediated inflammatory response with minimal side effects ^21^. The ability of Tocilizumab to calm the cytokine storm and halt the inflammatory processes that lead to poor outcomes in COVID-19 is still under investigation^13^. However, in our small, observational case-control study, there appears to be a clear survival benefit for the sickest patients.

Prior studies suggests COVID-19 patients treated with Tocilizumab show improvement in physiological markers such as fevers, oxygen requirements, CRP, lymphopenia and imaging abnormalities^9^. To our knowledge, our report is the first to show an improvement in clinically important outcome of mortality in COVID-19 patients treated with Tocilizumab. Further follow-up to determine the longer-term benefits of Tocilizumab after COVID-19 acute illness is required to understand the full benefit.

Our study suggests that IL-6R receptor inhibition may have a role to play in management of COVID-19 patients with severe respiratory complications. It remains to be seen if other IL-6 inhibitors are direct antagonists of IL-6 or inhibitors of downstream mediators such as Janus kinase (JAK) will have similar effects. In our patients, the therapy with Tocilizumab was administered relatively late in the course of the disease after the onset of ARDS as shown by the fact that more patients who received therapy were on mechanical ventilation or had higher oxygen requirements. It is possible that earlier administration of the drug before the onset of the ARDS in selected patients with high inflammatory markers such as CRP, D-dimer and ferritin would result in better outcomes.

Our study has several limitations. First, the sample size is small and includes selection bias. We addressed the selection bias by using inverse probability weighting in our modeling. Treatment was determined in the clinical setting and data were analyzed retrospectively. This is not as robust as a randomized controlled trial design. Second, our sample of patients was missing laboratory values that may have informed us otherwise about our models. For example, though many elements in our univariate analysis informed us that the patients in the Tocilizumab group were sicker than the untreated group, the average SOFA scores between the two groups was not statistically significant presumably due to presence of missing variables.

## Conclusions

This study demonstrates the potential effectiveness of Tocilizumab in a real-world setting for treatment of hospitalized patients with severe and life threatening COVID-19 disease. In the acute setting, Tocilizumab may be an option to improve survival in the aforementioned subgroup of patients with COVID-19 infection. Further study, with larger samples and randomization are needed to determine the efficacy of Tocilizumab in patients hospitalized with COVID-19 disease.

## Data Availability

Data are confidential and protected but may be made available to reviewers as de-idenfified, aggregate data.

